# Science or Advocacy? The Global Rise of Policy Claims in Population Health Research (1990-2024)

**DOI:** 10.1101/2025.11.13.25340175

**Authors:** David Bann, Mengyao Wang, Neil M Davies, Liam Wright, Meredith O’Connor, Emilie Courtin

**Affiliations:** Centre for Longitudinal Studies, University College London, UK; Department of Biostatistics, Yale University, US; LSE Health Inequalities Lab, Department of Health Policy, London School of Economic and Political Science, UK; Division of Psychiatry, University College London, UK; Department of Statistical Science, University College London, UK; Department of Public Health and Nursing, Norwegian University of Science and Technology, Norway; Melbourne Children’s LifeCourse Initiative, Murdoch Children’s Research Institute, Melbourne, Australia; The University of Melbourne, Department of Paediatrics, Melbourne, Australia; The University of Melbourne, Faculty of Education, Melbourne, Australia

## Abstract

Should original research routinely contain prominent policy claims, such as recommendations for policymakers or broad calls to action? Growing emphasis on “research impact” might be welcome but also have unintended consequences that include risks of overextrapolation, the blurring of roles between scientists and advocates, and potential erosion of scientific credibility. To inform this debate, we examined 45,807 abstracts from ten leading Epidemiology and Public Health journals (1990–2024). Using a large language model with human validation, we classified policy claims and mapped their prevalence by time, country, journal, field of study, and study design. Policy claims markedly increased in frequency from 17.6% in 1990-1999 to 35.8% in 2020-2024, with wide variation across countries (>40% in Italy and Australia vs <19% in Norway and Japan, 2020–2024) and journals (>60% in some vs <6% in others). Keywords linked to higher claim rates differed by topic and time: some corresponded to topics with clear causal evidence (“tobacco”), others to topics with more complex causal evidence and notable researcher advocacy (“health inequalities” and “COVID-19”). Claims were most common in qualitative or cross-sectional studies, and less common in cohort, quasi-experimental, or experimental studies. We argue that these patterns reflect a research culture increasingly oriented toward claiming policy relevance—and incentives that encourage attaching claims to single studies. Our findings raise questions about how scientists and journals balance evidence, advocacy, and credibility. Ensuring that policy claims remain commensurate with evidence will be central to build trust as policy impact continues to be incentivised. We discuss alternative ways for researchers and publishers to engage meaningfully with policy beyond attaching claims to individual studies, and share our data and scripts to catalyse further work in this area.

## Introduction

High-quality evidence is required to inform policymaking. This recognition, as well as the legitimate desire to ensure that research funding is used effectively, has led to researchers being increasingly expected to have ‘impact’ alongside conducting world-class research. Impact—on policy, society, or the economy—is now a key metric by which universities are awarded funding (e.g., in the UK^1^ and Australia^2^ since 2009-11). It also informs how researchers are rewarded such as via career progression^3^, personal awards^4^ ^5^, or grants (e.g., NSF in the USA^6^). This has been termed the ‘impact agenda’. There are many competing definitions and frameworks to assess the impact of research on policy.^7^ For example, impact in the UK is defined as “the demonstrable contribution that excellent research makes to society and the economy”.^8^

However, concerns have arisen about how the ‘impact agenda’ is operationalised (e.g., in research policy or funding), and the incentive structures it entails.^1^ ^9^ These incentives may have unintended consequences, including operating at cross-purposes with the more traditional aim of academic research: the dispassionate pursuit of truth.^10^ Researchers, who often lack formal training in policy analysis and operate in highly competitive environments, are increasingly rewarded for showing policy relevance. This, in turn, might encourage ‘bold’ or exaggerated impact claims. Indeed, over recent decades, researchers have increasingly used ‘hyped’ language in their research papers,^11^ funding applications,^12^ and impact reports.^13^ ^14^

One possible consequence of this trend is the risk of making policy recommendations that extend beyond what a single study, or even a particular study design, can credibly support.^15^ Such overstatements can mislead policymakers and the public about the strength and totality of evidence on a given issue, potentially leading to ineffective policy decisions and a broader erosion of trust in both scientists and public institutions. Beyond these concerns, the ‘impact agenda’ raises additional issues, including potential threats to academic freedom (as research priorities shift toward incentivised short-term policy goals), the transformation of scientists from seekers of knowledge into policy advocates, and the increasing bureaucratic burden of demonstrating impact.^16^

There has been little empirical investigation into the prevalence and evolution of policy claims in empirical research papers. A 1999 paper that sampled three US-based Public Health/Epidemiology journals found that 24% of papers made policy recommendations.^17^ This differed by journal; *Epidemiology ,* a journal with an explicit editorial policy to avoid policy recommendations in research articles, made the fewest claims (8%).^18^ However, this study precedes the rise of the ‘impact agenda’ and does not track changes across time or location (country). It is also limited by the small number of journals, which restricts our understanding of how policy claims have evolved over time and across countries, topics, and research designs.

We thus analysed policy claims in published abstracts from Epidemiology and Public Health journals spanning 1990 to 2024. Research in these fields might be particularly vulnerable to the unintended consequences of the ‘impact agenda’.^15,19^ We focus on abstracts of original research articles since 1) they are freely accessible to both policymakers and the public (unlike full texts), and 2) their length makes them suitable for large-scale algorithmic analyses.

Our aim is descriptive: to quantify trends in policy claims within the impact agenda. We do not assess the validity of individual claims. Our contribution is also methodological: to systematically map out policy claims across articles, we employed a frontier open-source large language model (LLM), which excels in understanding language differences,^20,21^ and allowed us to increase scale without loss of reliability.

## Methods

### Journal identification and abstract/metadata retrieval

We included multiple established or newly established Epidemiology and Public Health journals which publish original empirical research. We expanded upon the three journals included in Jackson et al^17^ 1999 (which used manual evaluation) by including non-US focused journals (e.g., European Journal of Epidemiology / European Journal of Public Health / Lancet Public Health). Given the lack of an agreed metric for journal quality or influence, author judgement and discussion informed this choice, resulting in 10 journals (see Table 1).

**Table 1.** Characteristics of Epidemiology/Public Health abstracts: policy claims by year, journal, and country.

|  | 1990-1999 | 2000-2009 | 2010-2019 | 2020-2024 | All years |
| --- | --- | --- | --- | --- | --- |
| **Number of articles, n** | 10436 | 12529 | 15464 | 7378 | 45807 |
| **Number of unique journals, n** | 9 | 9 | 10 | 10 | 10 |
| **Words in abstracts, mean (sd)** | 205.5 (67.5) | 215.9 (55.2) | 221.0 (52.0) | 238.4 (57.4) | 218.9 (58.5) |
| **Papers with keywords included, %** | 49.4 | 60.6 | 45.1 | 57.8 | 52.4 |
| **Countries of corresponding author, n** | 112 | 118 | 121 | 104 | 153 |
| **Policy claims, n (%)** | 1841 (17.6) | 2857 (22.8) | 4394 (28.4) | 2643 (35.8) | 11735 (25.6) |
| **Journals** |  |  |  |  |  |
| Epidemiology | 1.2 | 4.1 | 4.5 | 5.3 | 3.7 |
| European Journal of Epidemiology | 19.4 | 15.3 | 10.0 | 14.3 | 14.4 |
| American Journal of Epidemiology | 8.2 | 8.6 | 11.1 | 15.6 | 9.9 |
| International Journal of Epidemiology | 15.3 | 20.0 | 16.4 | 20.0 | 17.4 |
| Journal Of Epidemiology and Community Health | 20.8 | 25.9 | 30.5 | 41.1 | 28.8 |
| European Journal of Public Health | 28.1 | 33.5 | 40.4 | 45.2 | 38.7 |
| Preventive Medicine | 25.1 | 30.3 | 38.1 | 45.2 | 35.7 |
| American Journal of Public Health | 24.5 | 33.7 | 38.6 | 46.2 | 33.7 |
| American Journal of Preventive Medicine | 31.3 | 37.2 | 35.9 | 47.2 | 38.1 |
| The Lancet Public Health | N/A | N/A | 59.1 | 64.1 | 62.5 |
| **Countries** |  |  |  |  |  |
| Norway | 8.5 | 6.3 | 14.5 | 16.9 | 10.8 |
| Japan | 9.0 | 17.3 | 17.6 | 18.5 | 16.0 |
| Denmark | 12.8 | 9.1 | 11.3 | 24.9 | 13.4 |
| Sweden | 14.0 | 15.2 | 21.9 | 27.4 | 18.7 |
| Germany | 19.5 | 17.5 | 16.5 | 28.9 | 19.5 |
| Finland | 10.6 | 18.7 | 25.8 | 30.5 | 20.2 |
| China | 11.4 | 16.8 | 23.3 | 33.0 | 25.8 |
| Netherlands | 14.6 | 22.1 | 25.1 | 34.2 | 22.8 |
| United Kingdom | 16.6 | 23.1 | 25.5 | 34.3 | 24.7 |
| Spain | 16.2 | 24.6 | 26.8 | 34.4 | 25.3 |
| France | 17.0 | 19.9 | 25.3 | 34.5 | 23.5 |
| Canada | 16.9 | 23.1 | 29.9 | 34.8 | 26.9 |
| United States | 18.2 | 24.1 | 30.5 | 37.6 | 26.9 |
| Australia | 23.6 | 26.4 | 29.7 | 40.1 | 29.7 |
| Italy | 14.9 | 19.7 | 36.0 | 43.5 | 23.7 |

Focusing on articles published from 1990 to 2024 to capture periods before and during the impact agenda, we downloaded all abstracts and relevant metadata (e.g., year, keywords, citation count, country of corresponding author) using the Scopus Application Programming Interface (API^22^. Since our goal was to examine original research articles, we extracted only entries listed as “research articles”. We filtered further based on title and keyword fields to remove systematic reviews/meta-analyses or commentaries (n=50,533 records screened; n=4,725 records were excluded; One additional record lacked an LLM policy-claim label and was excluded from labelled analyses; analytical sample n=45,807). See Supplementary Table 1 for details of our sample selection.

### Defining policy claims using large language models (LLMs)

We employed large language models (LLMs) to identify policy claims in abstracts. Because LLMs excel at semantic understanding, we reliably detected policy recommendations at scale. These recommendations are often expressed through varied language patterns and use contextual nuances that can evade simple pattern-matching techniques.

We used the Deepseek v3.1 LLM, a frontier-level open-weight model.^21^ We chose a temperature of 0.1 (temperature is a measure of variability in model outputs; values range from 0 to 1.5 with values closer to 0 ensuring greater determinism in results at the expense of model flexibility). For each abstract, the LLM was prompted to identify explicit or implicit policy recommendations. Prompt engineering principles^23,24^ were used, which have been found to increase LLM performance in general (e.g., specifying LLM role and giving examples); the specific prompt used is shown in Supplementary Table 2. A statement within an abstract was defined as a policy claim if it directly suggested policy attention or calls for action, regulation, or intervention. This call to action could either be specific (e.g., “States should ban X”) or vague in nature (e.g., “This has implications for policy…”). Furthermore, the claim could be directed at a specific group, such as governments or public health organisations, or be more general (e.g., “future policies should”). Finally, to distinguish it from background information, the statement needed to appear in the abstract’s concluding sentences. Statements were explicitly not considered policy claims if they were a suggestion for future research, a finding or statement of fact with no call to action, or a background statement used for motivation.

#### Validation of LLM classification

To validate the classification approach, we first assessed reliability across three repeated runs on a random sample of 200 abstracts with random seeds not specified; this demonstrated almost perfect run-to-run reliability (Cohen’s κ=0.94−0.97, N=200). We then compared with human classification. As a baseline, inter-rater reliability between the two human lead authors (DB and EC) was also very high (κ=0.83, N=98). For context, a recent review of concordance in systematic review abstract screening in human raters reported κ=0.82.^25^ When compared with human classifiers, there was very high concordance with the LLM: with rater MW (κ=0.84, N=98), DB (κ=0.73, N=98), and EC (κ=0.71, N=100; EC additionally reviewed 10 additional abstracts where LLM policy claim=true: κ=0.74, N=110). Across all human-LLM comparisons, raw per cent agreement was also very high, ranging from 88% to 94%.

The cost and time to process such a large number of abstracts are dependent on the LLM compute / API costs; for the Deepseek API, for example, the analysis incurred ∼$3 and ∼10 hours of processing time. Since Deepseek is open-weight, the model can be run on local hardware with sufficiently high RAM.

### Analytical strategy

#### Policy claims by time, country and journal

We calculated the fraction of abstracts by classification status: overall, and then by year, country, and journal. We first did this in absolute terms (# per year, cumulatively), since the cumulative total of papers that make policy claims may be impactful for evidence translation. We next estimated the claims made in relative (percentage) terms overall and in each subgroup by year. To avoid excess variation over time by focusing on single years, we report statistics using categories (1990-1999, 2000-2009, 2010-2019, 2020-2024).

#### Comparison of study designs

To understand which study designs were used in abstracts which made policy claims, we classified study designs using the recorded keywords (e.g., either “cross-sectional studies”) and title/abstract fields (e.g., “a cross-sectional study”). We classified studies as experimental, quasi-experimental, cohort, cross-sectional, ecological/time-series, case-control, or qualitative. This resulted in a study design being assigned for 8,715 abstracts.

#### Comparison of research topics

To understand whether policy claims differed by research topic, we compared the keywords listed in abstract metadata by policy classification status. We described the percentage of policy claims amongst the most commonly reported keywords and amongst those with the highest rates of policy claims.

To understand the nature of the policy claims made in the abstracts, we randomly selected 100 articles for which we manually checked the content of the claims and classified it as either: (1) broad call for action; (2) further investment in existing policies or intervention; (3) call for new policy direction; (4) focus on specific subgroups/potential new beneficiaries of the policy or intervention; (5) new evidence for policy-making in the area of interest; (6) methods recommendations (7) extensions of existing policies.

#### Additional and sensitivity analyses

To examine if differences in claims were driven by differences in publication of non-empirical research, we conducted sensitivity analyses excluding abstracts with the keywords “epidemiologic methods” and “simulation”. We also examined trends using an alternative approach: using regular expressions to classify bold policy claims rather than an LLM (e.g., “policymakers should”, “government must”). While our piloting work suggested this led to a vast underestimation of policy claims owing to differing wording (kappa in the sample reviewed by humans was 0% since it did not capture any policy claims in the n=100 reviewed by humans), the regular expression approach may still be helpful to triangulate the main findings, if the degree of underestimation is consistent across time.

## Results

### Policy claims by time and country

Across the study period (1990-2024), 25.6% of abstracts were classified as making a policy claim. This increased continuously over time: from 17.6% in 1990-1999 to 35.8% in 2020-2024 (Table 1 and Figure 1).

**Figure 1.**
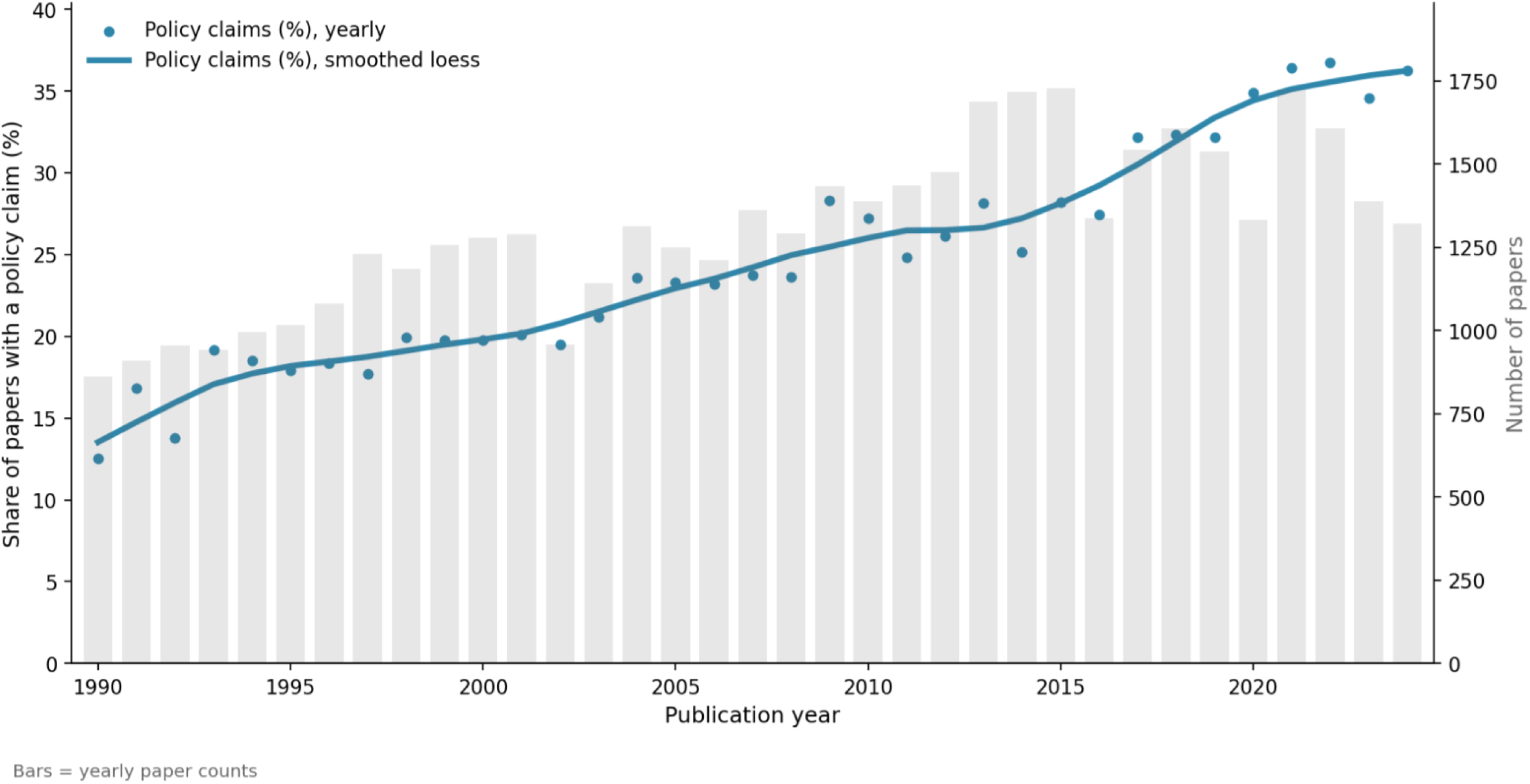
Share of abstracts with policy claims in Epidemiology/Public Health Abstracts, 1990–2024.

The prevalence and time trends of policy claims markedly differed by country (Table 1 and Figure 2; Supplementary Figure 1). For example, amongst the 15 countries which published the most papers, Italy (43.5%) and Australia (40.1%) had the highest prevalence in 2020-2024, with Japan (18.5%) and Norway (16.9%) the lowest. Italy had the largest increase from 1990-1999 to 2020-2024 (14.9% to 43.5%) and Norway had the lowest (8.5% to 16.9%).

**Figure 2.**
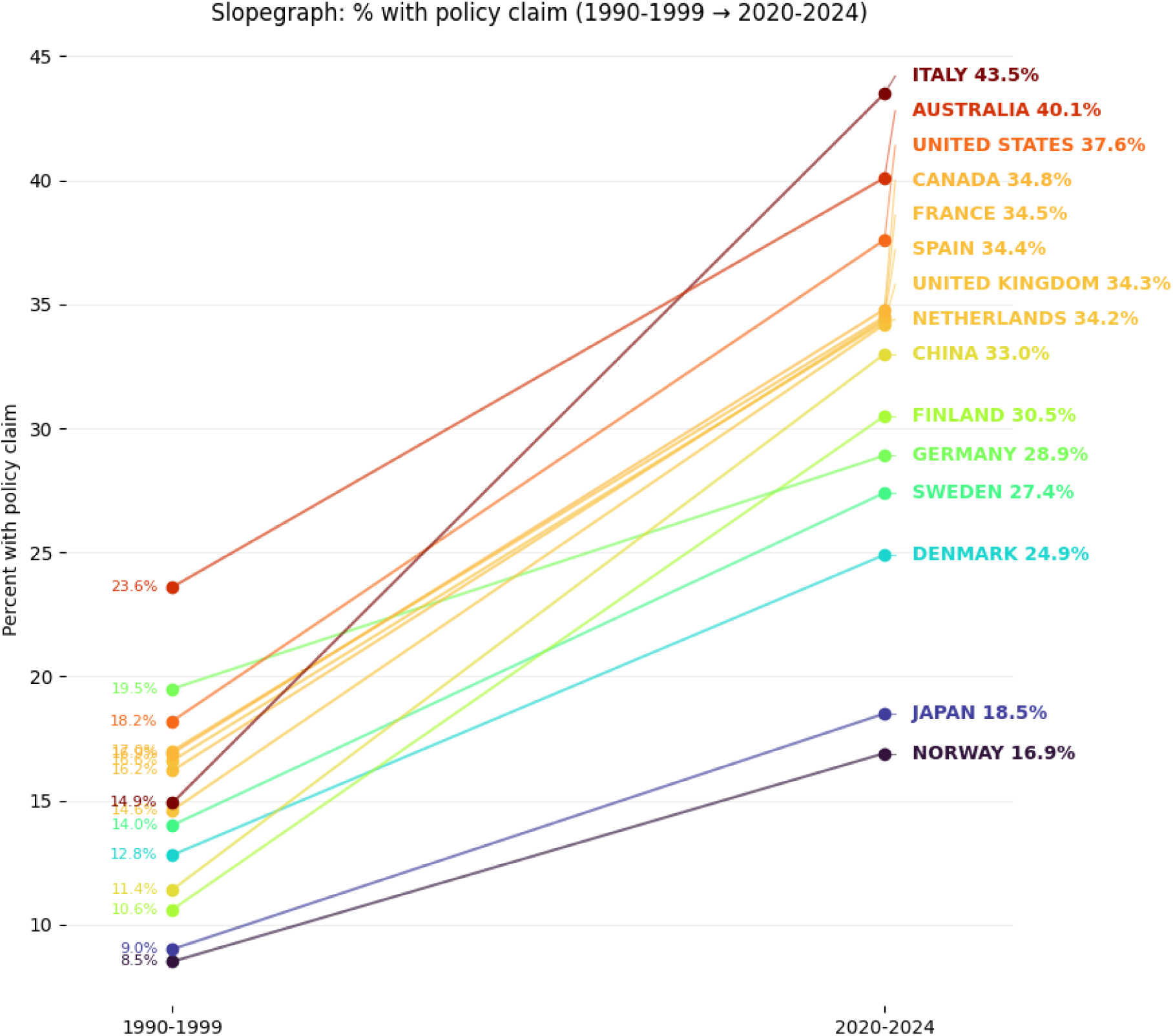
Policy claims by country, 1990–2024.

### Policy claims by journal

There was a striking difference in the frequency of policy claims made by journal: an over 16-fold difference, for example, comparing Epidemiology (3.7%) with Lancet Public Health (62.5%); Table 1. We also find clear heterogeneity in the change across time: for example, the Journal of Epidemiology and Public Health increased from 20.8% to 41.1%, while claims made in the European Journal of Epidemiology declined from 19.4% to 14.3% (Figure 3 and Table 1). In absolute and relative terms, public health-focused journals had higher claim rates than Epidemiology-focused journals (Figure 3).

**Figure 3.**
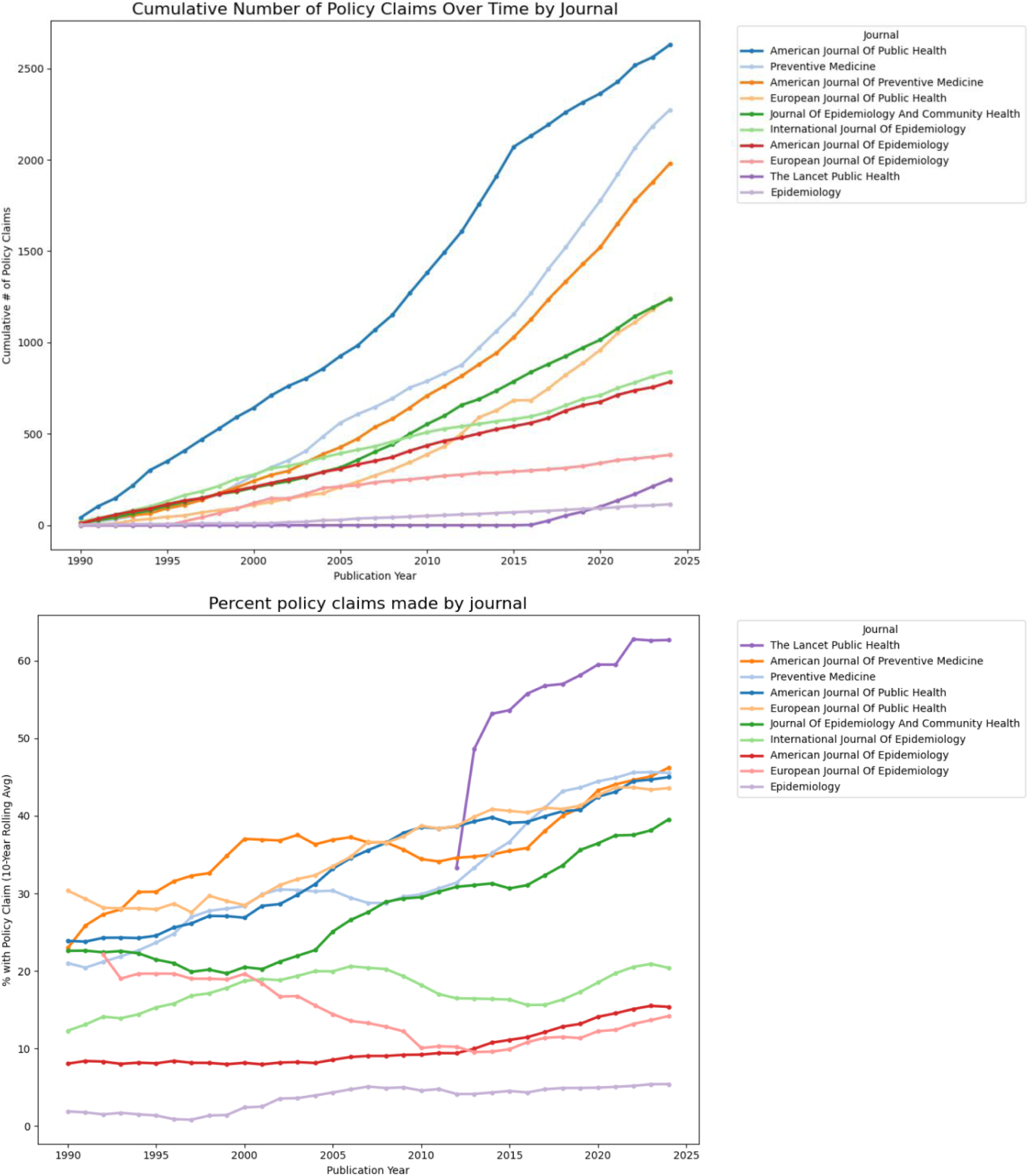
Policy claims by journal: in absolute terms (top) and relative percentage terms (bottom)

### Policy claims by study design

Amongst the subset of papers which could be algorithmically identified as having a clear study design, policy claims were most frequent in qualitative studies (48.39%), followed by cross-sectional studies (38.43%); Figure 4. Experimental (e.g., RCTs) and case-control studies had the lowest proportion of policy claims. The unexpectedly higher fraction of policy claims in qualitative studies was confirmed after manually validating the study design; removing mixed-methods or ambiguous cases yielded a 49.3% policy claim rate (n=73).

**Figure 4.**
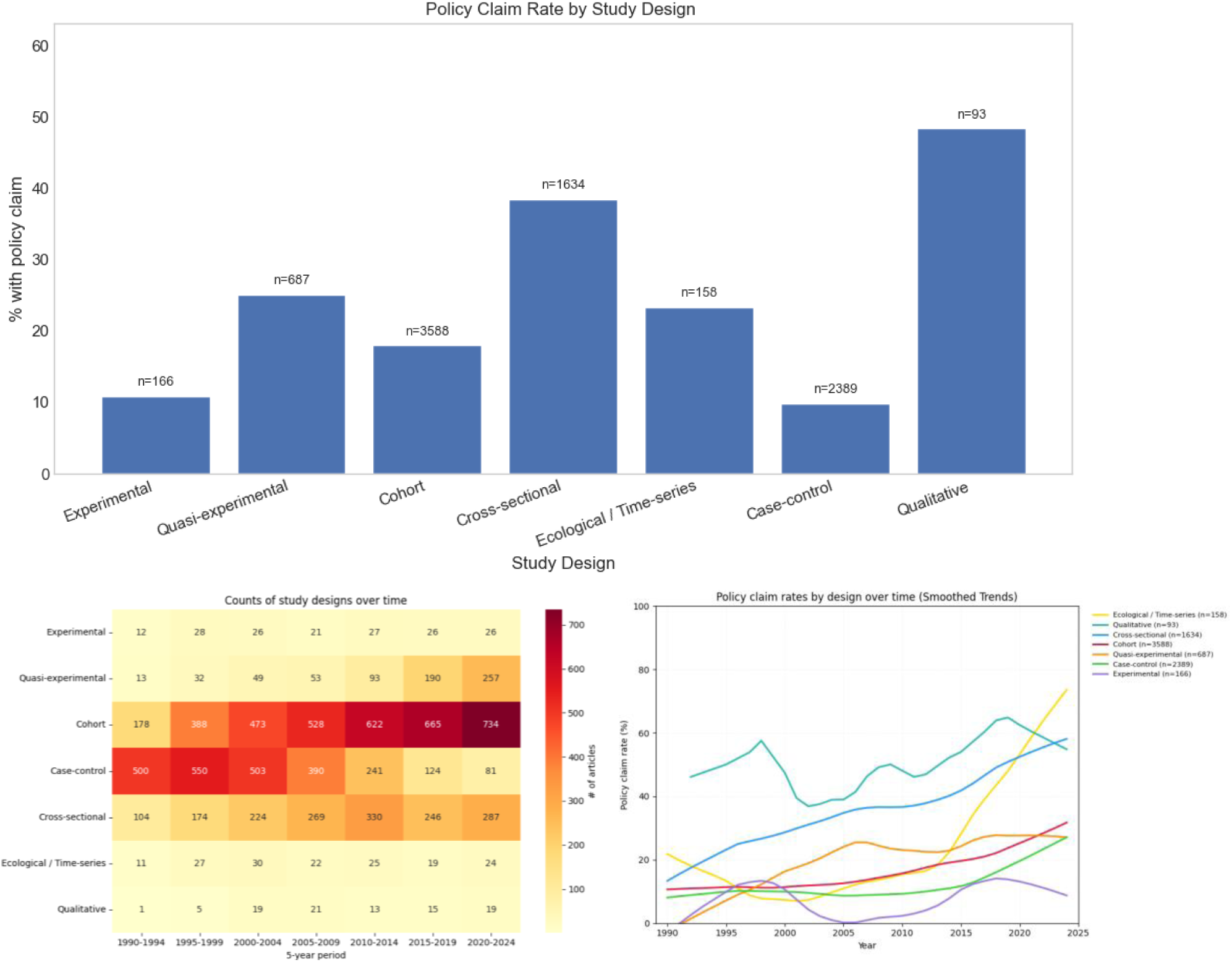
Policy claims rates by study design (upper and right panels), and tabulations of study design by year (left)

**Figure 5.**
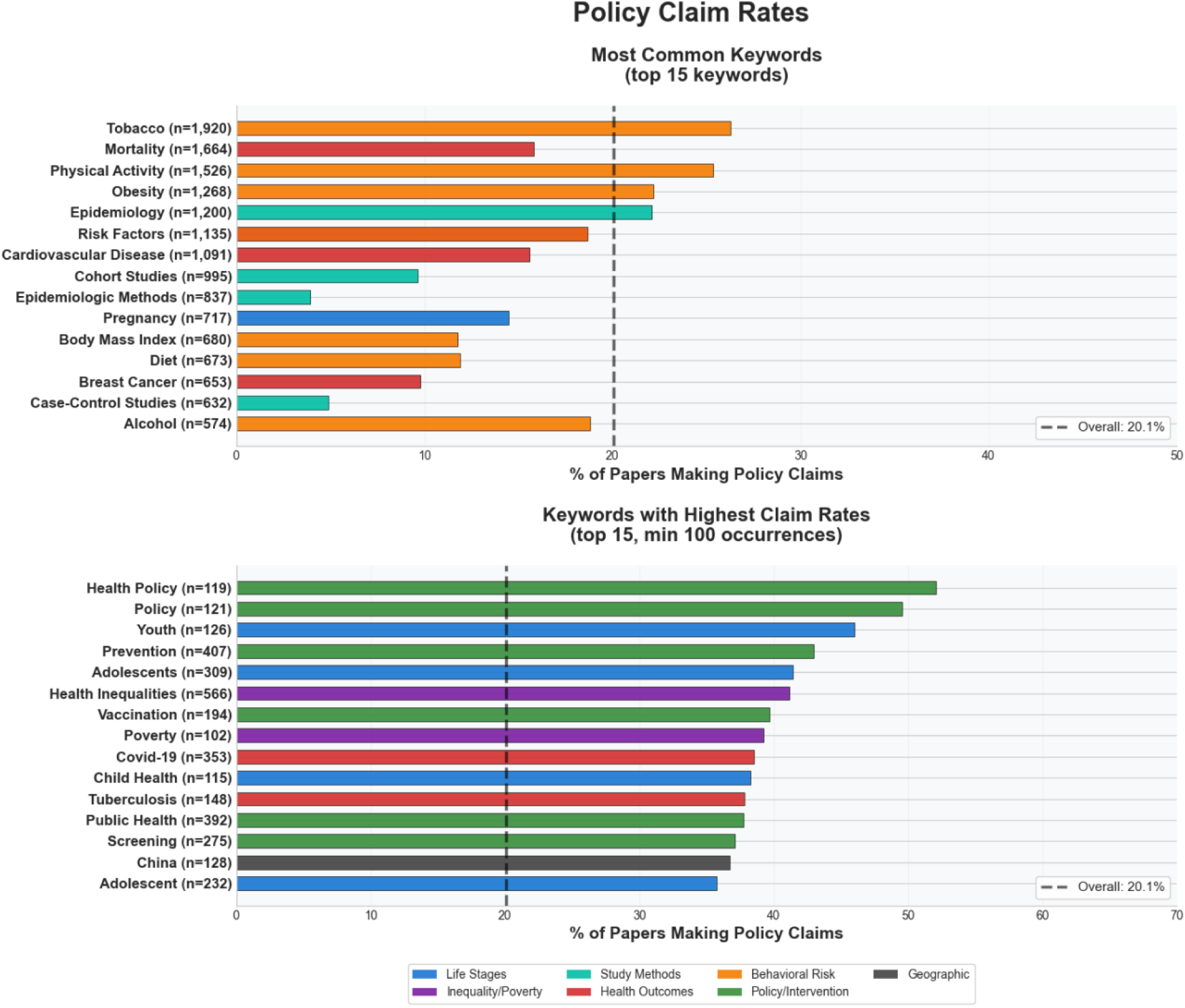
Policy claims by subject (keywords).

### The characteristics of claims: keywords and types of claims

The rates of policy claims differed by article keywords. Policy claims were, for example, higher than average for fields with clear causal evidence of harm (e.g., tobacco, 26.3%) and for topics with notable researcher advocacy, e.g., health inequalities (41.2%), poverty (39.2%), and COVID-19 (38.5%). Keywords with the highest prevalence of policy claims differed across time (e.g., research on adolescence and smoking cessation featured prominently in 2000-2009 vs mental health and health inequalities in 2020-2024); Supplementary Figure 2.

A manual screen identified multiple types of policy claim (n=100); Supplementary Table 3. After excluding 3 claims where it was unclear if a policy claim was made: 38% highlight alternative policy approaches (e.g., wage-setting policies for food insecurity), 27% recommend that policies should differentially target or tailor interventions to sub populations (e.g., older migrants), and 25% flag their empirical findings as relevant or important for policy audiences (e.g., depression’s link with employment). The remaining include calls for action (5%), calls for further investment in existing policies (1%), and extensions of existing policies (1%).

### Additional and sensitivity analyses

The substantial rise in policy claims was also found when 1) removing papers with “epidemiologic methods” or “simulation” in keywords (2.3%; Supplementary Table 4); and 2) when using regular expressions for bold policy claims rather than an LLM to classify policy claims (e.g., “policymakers should”, “government must”; Supplementary Figure 3).

## Discussion

We observed a marked increase in policy claims in published Epidemiology and Public Health abstracts from 1990 to 2024. This coincides with the rise of the impact agenda, with particularly large increases in countries notably affected, such as the UK, Australia, and the USA.^26^ ^27^ The sharp rise in policy claims in Italy (19.7% in 2000-2009 to 36.0% in 2010-2019), while not anticipated a priori, is consistent with the 2010 introduction of metrics in Italian academia, which reportedly led to undesired consequences (increasing self-citation rates).^28^

The substantial variation in policy claims suggests that scientific reporting practices are sensitive to external pressures; here, a research culture is increasingly incentivised to demonstrate policy relevance.^1,9,16,27^ Differences across journals also indicate the importance of editorial policy: for example, Epidemiology, which explicitly restricts policy claims to commentaries, had the lowest rate, whereas journals that explicitly encourage policy relevance had rates up to 16-fold higher.

Whether this large increase is supported (reflecting shifts in the research questions chosen) or not supported (reflecting overextrapolation or hype) by the studies the abstracts summarise cannot be precisely determined using our data alone. Discerning overextrapolation requires deep contextual understanding of each abstract’s specific research question. The claims we identified likely span a spectrum of defensibility. Some are justified within the narrow context of their study findings; others are only defensible when interpreted in a broader sense, for example, when the policy claim refers to an accumulated body of evidence; others are broad statements that motivate further policy interest in a subject. While it is generally advisable that single research studies should not lead to direct policy recommendations,^15,19^ exceptions exist in fields where the evidence base is fundamentally constrained.

Several lines of reasoning, however, raise questions about a potential increase in overextrapolation or in the use of bold or ‘hyped’ policy claims. First, the increase in policy claims was not accompanied by a marked rise in claims from studies using rigorous causal-inference designs (quasi- and experimental studies; rates were highest for qualitative studies). This would have indicated that claims were increasingly supported by stronger methodological foundations and approaches explicitly designed to assess policy impacts. The absence of this pattern suggests that incentives to signal policy relevance may have outpaced methodological rigour. Second, policy claims were high in topics marked by causal uncertainty, where overextrapolation concerns have been explicitly raised, such as research on health inequality^29^ or COVID-19.^30^ Third, increases in policy claims were also observed using an alternative specification that focused on bold terms (e.g., “policymakers should”). Fourth, such increases are anticipated given the competitive landscape researchers and journals operate in: previous studies report increases in promotional or ‘hyped’ language when communicating scientific importance in policy impact reports,^13^ ^14^ papers,^11^ and grant submissions.^12^

The potential for a cumulative body of overstated claims to erode scientific credibility is concerning, even if our estimates are upwardly biased. For example, if only 10% of abstracts classified as making a policy claim used hype or overextrapolation, this would still be 1,174 papers making such claims.

A related practical issue concerns where and how policy discussion should be presented to optimally contribute to evidence-based policy. The routine inclusion of policy claims in research abstracts may in part be attributable to the limitations of conventional publishing practices—the legacies of 20th-century paper constraints applied to electronic content. Abstract word limits leave little space for the broader context that authors may implicitly intend (e.g., “the collective body of evidence suggests…”). Even in journals that include “evidence summary” or “policy relevance” sections, it is often unclear whether abstract claims refer to the single study or the wider literature. Space within full articles is also limited, restricting opportunities for detailed, evidence-based deliberation of policy options. Alternatives include placing detailed policy discussions in appendices or publishing separate, dedicated policy pieces.^18^

By sharing our methods and code, we aim to encourage further research on the nature, determinants, and consequences of policy communication, and to demonstrate the utility of large language models for computational metascience. Although copyright restrictions prevent redistribution of the original abstracts obtained from Scopus, we make available publicly available metadata along with our LLM classifications. Similar analyses could readily be extended to other disciplines. Economics and other quantitative social sciences, for example, also research population health topics, and economics underwent a “credibility revolution”^31,32^ marked by a focus on causal identification.

### Strengths and limitations

Our analysis benefits from the use of large language models (LLMs), enabling abstract review at scale—the full population of papers rather than a small sample. This enhances power and representativeness and provides a new resource for future research. Concordance between human and LLM reviewers was high, though all human reviewers^14^ inevitably make errors. While all such computational approaches may contain errors, such errors would need to vary substantially across subgroups (e.g., journal and year) to meaningfully alter our main findings. Triangulation using a regular-expression approach produced qualitatively similar findings, though it captured substantially fewer policy claims.

Limitations include the reliance on abstracts, chosen for their accessibility to policy/public audiences and suitability for algorithmic analysis. A comprehensive understanding of impact communication would require examining the full set of researcher outputs, including full texts, press releases, conference presentations, and informal advocacy. Our use of Scopus to extract abstracts reflects both coverage and access constraints; while we piloted the open-access alternative OpenAlex, it lacked full access to copyrighted abstracts. Although we restricted ourselves to articles labelled as “original research,” some non-empirical papers may remain. Reassuringly, sensitivity analyses with additional exclusions (e.g., removing abstracts containing methodological keywords) yielded similar findings. Future research could compare policy claims across article types, such as editorials, original research, and reviews, and expand the corpus of included journals (field-specific and general).

## Conclusion

We observed a marked increase in policy claims over time, with notable variation across countries, journals, fields of study, and study design. These patterns occurred in the context of a shifting research culture increasingly oriented toward policy relevance—and incentives that encourage attaching claims to single studies. Our findings raise questions about how scientists and journals balance evidence, advocacy, and credibility. We encourage greater discussion of such issues, and make our data and code freely available to help catalyse further work.

## Code availability

Analytical code to reproduce the findings, and the key datasets used, are available at: https://github.com/dbann/policyclaims.

## Data Availability

Data are available at https://github.com/dbann/policyclaims

## Acknowledgments

We thank Majeic Piechocki for research assistance with an earlier iteration of this work.

## Contributions

Wrote the first draft: DB. Contributed to revising the text and interpretation: all authors. Conducted main analyses: DB. Conducted code review and contributed to analyses: MW. Contributed to manual screening: MW, EC, DB.

