## Supplementary for "Science or Advocacy? The Global Rise of Policy Claims in Population Health Research (1990-2024)"

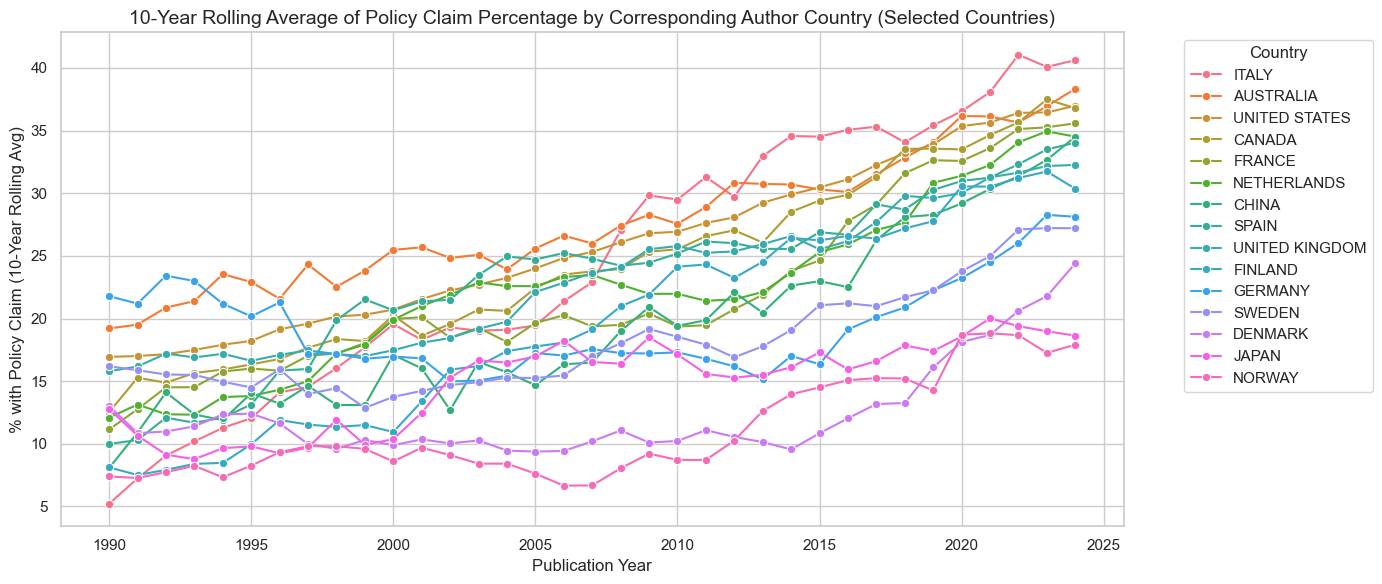

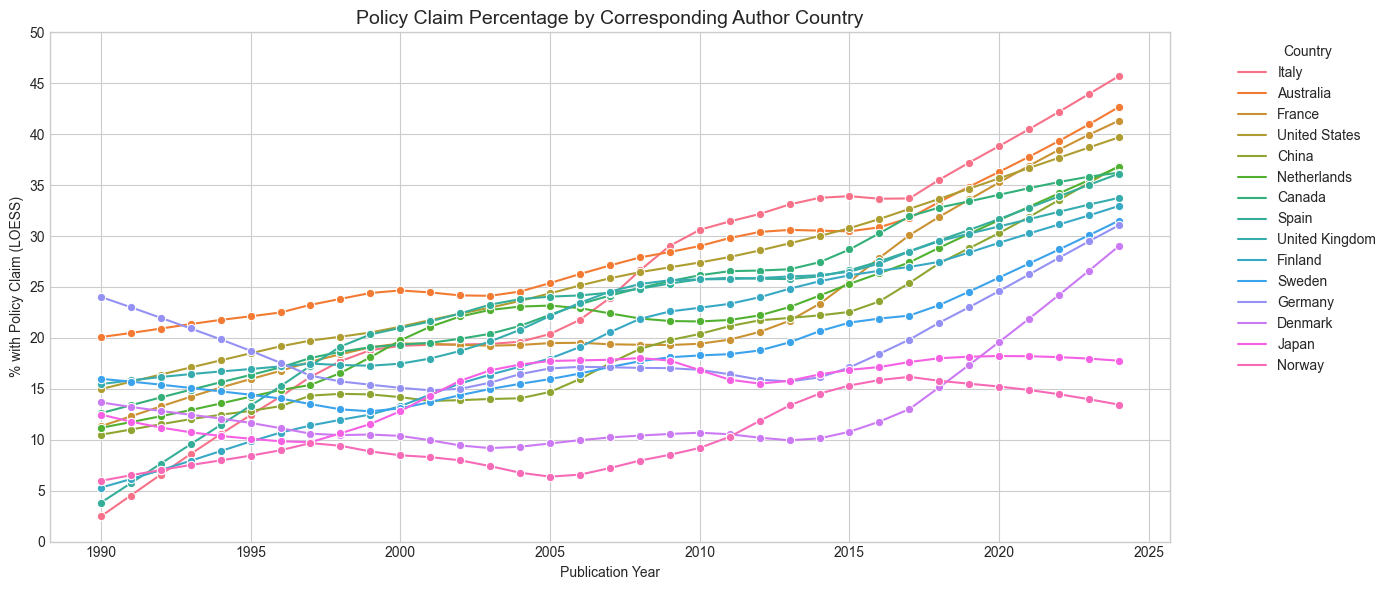

**Supplementary Figure 1. Policy claims by country, using rolling averages (top) or loess (bottom)**

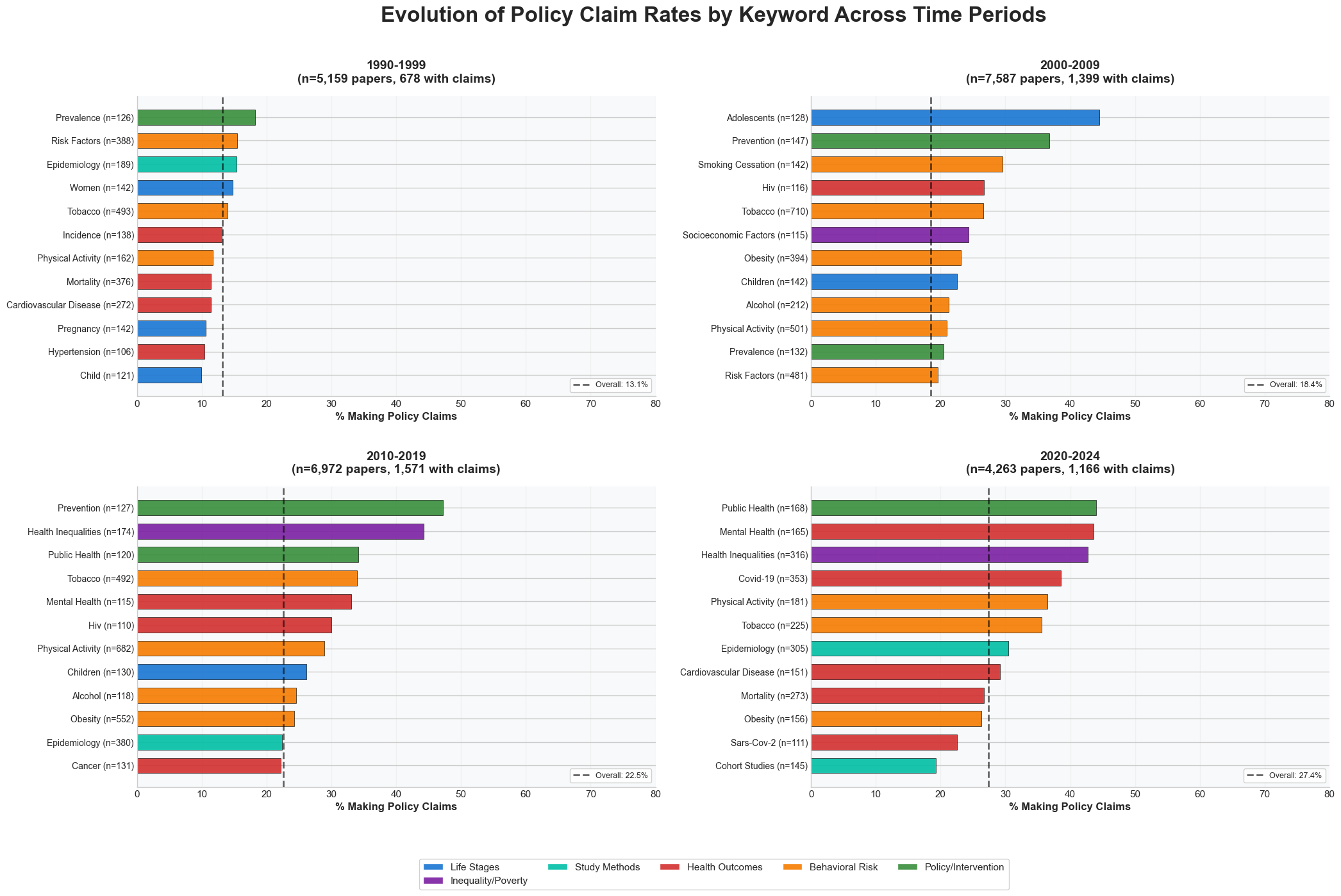

**Supplementary Figure 2. Policy claims by keyword/topic of study, by year**

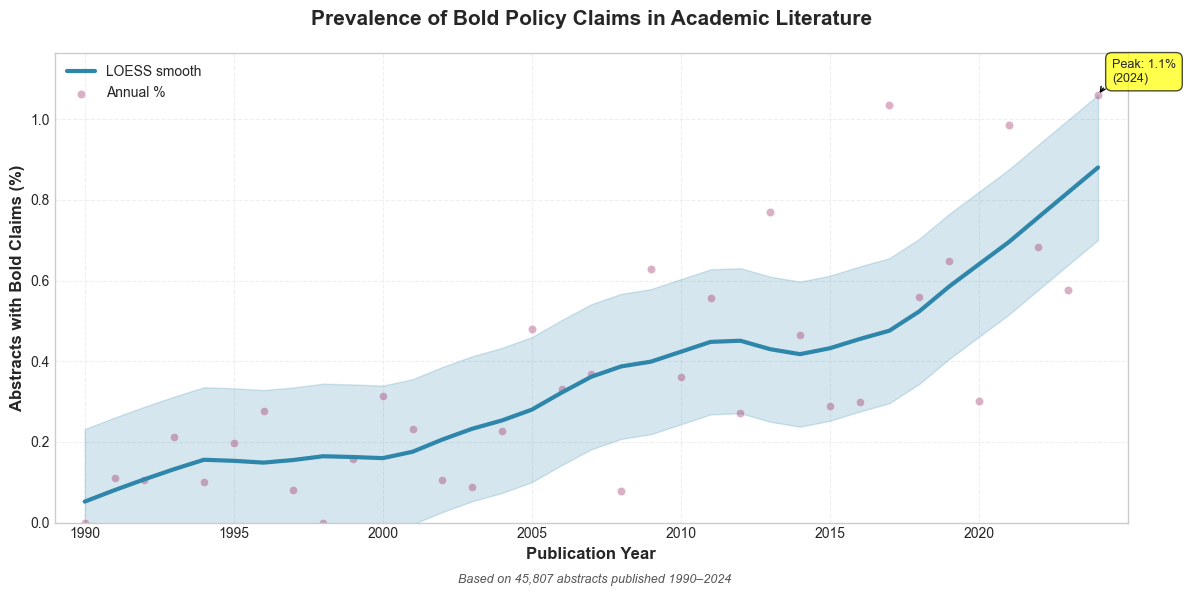

**Supplementary Figure 3. Policy claims using regular expressions**

**Supplementary Table 1. Journals Used.**

| Journal Name | Scopus ID | ISSN | Years | N (total) | N (analytical N: excluding missing abstracts, commentaries and review) |
| --- | --- | --- | --- | --- | --- |
| American Journal of Public Health | S168049282 | 0090-0036 | 1990-2024 | 8,770 | 7,807 |
| American Journal of Epidemiology | S170967050 | 0002-9262 | 1990-2024 | 8,433 | 7,926 |
| Preventive Medicine | S20040 | 0091-7435 | 1990-2024 | 6,655 | 6,368 |
| International Journal of Epidemiology | S27024 | 0300-5771 | 1990-2024 | 6.003 | 4,815 |
| American Journal of Preventive Medicine | S20224 | 0749-3797 | 1990-2024 | 5,710 | 5,188 |
| Journal of Epidemiology and Community Health | S156988948 | 0143-005X | 1990-2024 | 4,792 | 4,298 |
| Epidemiology | S15470582 | 1044-3983 | 1990-2024 | 3,437 | 3,122 |
| European Journal of Public Health | S4210220588 | 1101-1262 | 1991-2024 | 3,420 | 3,210 |
| European Journal of Epidemiology | S48690275 | 0393-2990 | 1996-2024 | 2,858 | 2,674 |
| The Lancet Public Health | S2764808104 | 2468-2667 | 2016-2024 | 455 | 400 |
| Total |  |  |  | 50,533 | 45,808 |

**Supplementary Table 2. Prompt used to large-language model**

LLM_PROMPT_TEMPLATE = """You are an expert policy analyst and academic researcher. Your sole function is to analyze a research abstract and determine if it makes a policy claim based on a strict set of rules and examples.

\## Rules for Classification

A statement **IS** a policy claim if:

- It directly suggests or calls for action, regulation, or intervention.

- It can be **vague** (eg, "This has implications for policy") or **specific** (e.g., "States should ban X").

- It may be directed at a specific body or group (e.g., governments, public health organizations, healthcare professionals) or be vague (eg, “future policies should”).

- It appears in the concluding sentences.

A statement **IS NOT** a policy claim if:

- It is a suggestion for **future research**.

- It is a finding or a statement of fact with **no call to action**.

- It is a background statement that **motivates the research** (usually at the start of the abstract).

\## Examples

ABSTRACT: "Our analysis of traffic data from 2020-2024 revealed that the new roundabout reduced accidents by 45%. These findings have significant implications for urban planning policy and should be considered by municipal transport authorities."

OUTPUT: {"policy_claim": true}

ABSTRACT: "We conducted a randomized controlled trial of a new diabetes drug. While the drug showed promise, there was no statistically significant improvement over existing treatments. Further investigation with a larger sample size is warranted to determine its efficacy."

OUTPUT: {"policy_claim": false}

ABSTRACT: "Our study shows a strong correlation between green space exposure and reduced symptoms of anxiety. To improve public health, municipal governments should enact zoning policies that mandate the inclusion of parks and green areas in all new housing developments."

OUTPUT: {"policy_claim": true}

ABSTRACT: "This paper reviews the historical literature concerning the United Kingdom's housing crisis. The data show that housing affordability has declined steadily since the 1980s across all regions, presenting a significant challenge for young adults."

OUTPUT: {"policy_claim": false}

ABSTRACT: "These results have important policy implications..."

OUTPUT: {"policy_claim": true}

\## Output Format

Your response MUST be a valid JSON object and nothing else. Follow this exact schema:

{"policy_claim": true/false}

\## Task

Now, analyze the following abstract based on all the rules and examples provided.

ABSTRACT: {abstract}

OUTPUT:"""

**Supplementary Table 3. 100 randomly selected abstracts, along with policy claim wording and manual categorisation of claim type.**

| **Doi** | **Policy Claim Wording Selection** | **Category of policy claim** |
| --- | --- | --- |
| 10.1016/j.amepre.2020.08.006 | These differences in the characteristics of registrants can help inform the tobacco control mass media purchasing strategies and may enable media efforts to target the specific subgroups of smokers in a better way. | Focus on specific subgroups |
| 10.1016/j.ypmed.2024.107944 | The analyses indicated that concurrent exposure to SB and OPA (odds ratio [OR] 1.36, 95% confidence interval [CI] 1.02–1.80) significantly raised the likelihood of increased hs-CRP, while SB without LPA (OR 1.26, 95% CI 1.11–1.41) or TPA (OR 1.19, 95% CI 1.08–1.31) significantly increased risk of higher hs-CRP, compared to their respective reference. The absence of non-occupational PA such as LPA or TPA combined with SB was associated with the increased hs-CRP risk, whereas OPA increased the risk when present concurrently with SB. Reducing SB and OPA while increasing LPA and TPA, is necessary to reduce inflammatory conditions. | Not a policy claim |
| 10.1016/j.amepre.2021.05.041 | The unadjusted incidence of maternal mortality in women with intellectual and developmental disabilities was 284 per 100,000 deliveries, nearly 4-fold higher than in women without intellectual and developmental disabilities (69 per 100,000 deliveries; risk ratio=4.07, 95% CI=2.04, 8.12), and the risk remained almost 3-fold higher after adjustment for sociodemographic characteristics (risk ratio=2.86, 95% CI=1.30, 6.29) and the expanded obstetric comorbidity index (risk ratio=2.30, 95% CI=1.05, 5.29). Conclusions: Women with intellectual and developmental disabilities are at increased risk of severe maternal morbidity and maternal mortality. These findings underscore the need for enhanced monitoring of the needs and maternal outcomes of women with intellectual and developmental disabilities in efforts to improve maternal health. | Focus on specific subgroups |
| 10.1136/jech-2020-216064 | We estimated there were an additional 4575 children with obesity (95% CI 1751 to 7399) and 9174 overweight or obese (95% CI 2689 to 15 660) compared with expected numbers had funding levels been maintained. Cuts to spending on Sure Start children’s centres were associated with increased childhood obesity. With deprived areas experiencing bigger spending cuts, reinvesting in these services may, alongside wider benefits for child development, contribute to reducing inequalities in childhood obesity. | Call for further investment in existing policy |
| 10.2105/AJPH.2021.306688 | Recommendations to promote structural equity are offered. | Call for action |
| 10.1016/j.ypmed.2021.106751 | Healthcare organizations have the opportunity to expand their lobbying on upstream SDOH policy issues to increase the impact of their SDOH strategy and further improve population health. | Call for action |
| 10.2105/AJPH.2021.306448 | Youth e-cigarette use patterns differed considerably across racial/ethnic groups, and tailored strategies to address disparities in e-cigarette use are needed. | Focus on specific subgroups |
| 10.1016/j.ypmed.2023.107680 | Key factors increasing the odds of knife-related victimization treated in EDs were homelessness, legal involvement, and substance use, particularly alcohol and stimulant use disorder. Somewhat surprisingly, mental health diagnosis was not associated with increased knife-related victimization. Although EDs are critical to treating knife-related victimization, they are also potentially key points to launch prevention for high-risk individuals to reduce subsequent violence stemming from escalation of interpersonal disputes. | Focus on specific subgroups |
| 10.1093/eurpub/ckab102 | This was greater than the sum of RDs for those with low MHC and non-smoker parent(s) [7% (2 14%)] plus those with high MHC and whose parent(s) smoked [11% (7 15%)]. There was limited effect measure modification by moderate or High Moderate MHC. Conclusion: Improving MHC to moderate levels may help reduce intergenerational transference of smoking. | Focus on specific subgroups |
| 10.1136/jech-2024-222293 | Policymakers should carefully evaluate the potential health consequences for specific demographics when introducing new welfare policies. | Focus on specific subgroups |
| 10.1016/j.ypmed.2020.106224 | Strategies are needed to help less active children to increase physical activity throughout childhood and adolescence to improve young adult health outcomes. | Focus on specific subgroups |
| 10.1007/s10654-022-00917-x | This epidemiologic study during the pandemic in professional musicians indicates no increased risk of SARS-CoV-2 infections in orchestra musicians and a trend towards increased risk in choir singers compared to controls. However, the exact routes of infection could not be validated. If appropriate hygiene concepts are adhered to, safe orchestra and choir activity appears possible in pandemic times. | Extension of existing policy |
| 10.1016/j.amepre.2019.09.017 | Conclusions: Low potassium intake in India warrants dietary policies promoting intake of potassium-rich foods to improve heart health. This approach may be more acceptable than programs focused on sodium reduction alone. | Alternative policy |
| 10.1016/j.ypmed.2023.107768 | Conclusion: Given the potential adverse effects of cannabis, prevention and harm reduction efforts should focus on groups at increasingly higher risk for use, including those with disabilities and kidney disease. | Focus on specific subgroups |
| 10.1016/j.amepre.2021.05.034 | Three quarters of caregivers for people with dementia reached by Area Agency on Aging staff were provided with information about relevant resources. Conclusions: The primary care liaison model is feasible, fosters ongoing interactions between primary care and Area Agencies on Aging, and connects older adults and their caregivers to relevant programs and services. Adoption of the primary care liaison model by other Area Agencies on Aging across the U.S. may help further the vision of optimized health and well-being of older adults. | Alternative policy |
| 10.1093/aje/kwad078 | Research funding agencies need to be engaged as mediators between committee needs and the research community to stimulate contributory research. Improved communication of research needs to the epidemiology community would be beneficial to researchers aspiring to have an impact and to those who use epidemiologic information to help guide policy decisions. | Call for action |
| 10.1016/j.amepre.2023.12.010 | Targeted interventions combining weight management and smoking cessation have been successful among the general population and could be adapted for pregnant individuals who smoke to facilitate cessation and healthy GWG in both urban and rural areas. | Focus on specific subgroups |
| 10.1093/eurpub/ckab080 | These results underline the importance of tailored preventive interventions in older migrants to detect and prevent these limitations at an early stage. | new evidence for policymaking |
| 10.1016/S2468-2667(20)30201-2 | A rebound in future workload could be imminent as COVID-19 restrictions ease and patients with undiagnosed conditions or delayed diagnosis present to primary and secondary health-care services. Such services should prioritise the diagnosis and treatment of these patients to mitigate potential indirect harms to protect public health. | alternative policy |
| 10.1016/j.amepre.2019.12.012 | Conclusions: Indicated or selective strategies are urgently needed to target micro-geographic locations with known increased risks, supplementing universal strategies applied to a broader community. | alternative policy |
| 10.1136/jech-2022-219729 | Policymakers considering implementing 20 mph speed limit interventions should consider the fidelity, context and scale of implementation. | new evidence for policymaking |
| 10.1016/j.amepre.2024.06.014 | Conclusions: Temperature extremes were associated with higher rates of missed primary care appointments. Individuals with chronic diseases were more likely to have missed appointments associated with extreme temperatures. The findings suggest the need for primary care physicians to explore different modes of care delivery to support vulnerable populations, such as making telemedicine during extreme weather events a viable and affordable option. | focus on specific subgroups |
| 10.1016/j.amepre.2021.08.020 | Clinical care for Veterans with military sexual trauma should consider elevated risk of opioid use disorder and high-risk opioid prescription. | alternative policy |
| 10.2105/AJPH.2020.306064 | The stark variation in DFLE and DLE across states highlights the large health inequalities present today across the United States, which have significant implications for individuals' well-being and US states' financial costs and medical care burden. | Not a policy claim |
| 10.1093/eurpub/ckae128 | Including plain packaging requirements in revising the European Union's legislative frameworks for tobacco control will help build progress towards a Tobacco-Free Europe without exacerbating smoking inequalities. | alternative policy |
| 10.1016/j.amepre.2023.09.002 | Conclusions: Approximately one in four U.S. adults aged <65 years is now recommended to receive PCV15 or PCV20, which highlights the need for providers to assess vaccination status, administer the vaccine, or refer patients as appropriate, as well as the need for tools to facilitate patient identification and vaccination. | alternative policy |
| 10.1016/j.ypmed.2021.106663 | More research is needed to disentangle the complex relationships between different neighbourhood built characteristics and specific types of sedentary behaviour. | focus on specific subgroups |
| 10.1016/j.amepre.2022.12.009 | Conclusions: To reduce the risk of child maltreatment and subsequent adverse childhood experiences, healthcare providers should screen parents for the presence of household challenges during both pregnancy and early childhood and connect patients to resources targeted at reducing those challenges and providing continuous familial support. | alternative policy |
| 10.1093/eurpub/ckaa077 | Our findings highlight the importance of an open and culturally sensitive attitude of the staff and the need to engage parents and community as a key to improve immigrant youths' accessibility to health care. | alternative policy |
| 10.1007/s10654-021-00766-0 | These findings provide evidence to support policy decision-making regarding which NPIs to implement to control the spread of the COVID-19 pandemic. | new evidence for policymaking |
| 10.1136/jech-2023-220750 | At the local level, in many European cities, time poverty could be reduced, among other interventions, by increasing affordable and good quality public services for the care of dependent persons. | alternative policy |
| 10.1016/j.amepre.2023.10.009 | Defining and understanding the interplay of these variables can guide policymaking and identify avenues to improve BCS for vulnerable or traditionally under-resourced populations. | focus on specific subgroups |
| 10.2105/AJPH.2021.306282 | These data highlight potential barriers to 7 key digital engagement behaviors that could be targeted for intervention. | new evidence for policymaking |
| 10.1093/eurpub/ckad022 | Conclusions: Appropriate screening and intervention programs are necessary for children with problem-drinking parents especially when exposure is severe but also at mild forms of exposure. | alternative policy |
| 10.2105/AJPH.2024.307804 | Policymakers need to be equipped with data to inform decisions about vaccine mandates in light of contextual factors and potential backlash affecting public health interventions. | Call for action |
| 10.1016/S2468-2667(22)00258-4 | Interpretation: Inviting eligible adults for lung health checks in areas of socioeconomic and ethnic diversity should achieve favourable participation in lung cancer screening overall, but inequalities by smoking, deprivation, and ethnicity persist. Funding: GRAIL. | new evidence for policymaking |
| 10.1016/j.amepre.2021.11.004 | Antimicrobial stewardship strategies are needed to improve prescribing by dentists. | alternative policy |
| 10.1016/j.ypmed.2024.107848 | Future campaigns should tailor messages to specific subgroups to broaden the reach (e.g., males), co-create materials with the target group, and give special attention to the contribution of metabolic/cardiovascular risk factors to dementia risk. | focus on specific subgroups |
| 10.1093/ije/dyab057 | Policies to support the effective isolation of cases from their household contacts could lower the level of household transmission. | alternative policy |
| 10.1136/jech-2019-213151 | In line with growing concerns over the potential consequences of austerity and associated policy measures, our findings suggest that these reform efforts pose a threat to the health of socioeconomically disadvantaged populations. | new evidence for policymaking |
| 10.1016/j.ypmed.2023.107679 | The urban-rural health disparities are significant and influenced by sociodemographic attributes, highlighting the importance of developing public health interventions to improve health outcomes in these populations. | focus on specific subgroups |
| 10.2105/AJPH.2020.305974 | Our policy proposal would allow historically marginalized community members to participate with dignity in the blood donation process without compromising blood donation and transfusion safety outcomes. (Am J Public Health. 2021;111:247-252. https://doi.org/10.2105/ AJPH.2020.305974). | focus on specific subgroups |
| 10.1016/j.ypmed.2022.106987 | These results necessitate public health efforts to ensure equitable and accessible healthcare as the COVID-19 pandemic continues. | new evidence for policymaking |
| 10.2105/AJPH.2020.305772 | Addressing social needs should be a priority of PWMIs to improve intervention adherence and reduce disparities in childhood obesity. | focus on specific subgroups |
| 10.2105/AJPH.2021.306589 | Fifty-two counties experienced decreases in ALS and CLS. Conclusions. Responding to trends in the gap between ALS and CLS at national and local levels is essential for the collective well-being of our nation, especially as we navigate and emerge from crisis. | Call for action |
| 10.1016/S2468-2667(23)00220-7 | These findings could be considered a baseline for monitoring the prevalence of clinically relevant depressive symptoms in Europe, and could inform policy for the development of preventive strategies for depression both at a country and European level. | new evidence for policymaking |
| 10.1016/j.ypmed.2020.106384 | Sexual minority-tailored interventions may be warranted to prevent tobacco product initiation. Worth exploring are the associations between sexual identity, tobacco marketing exposure, and friend(s)’ e-cigarette use. | focus on specific subgroups |
| 10.1093/aje/kwab067 | Blacks with higher levels of flourishing had a mortality rate that was not significantly different from that of Whites. However, Blacks, but not Whites, with low flourishing scores had a higher mortality rate. As such, health-promotion efforts focused on enhancing flourishing among Black populations may reduce the Black-White gap in mortalityrate. | alternative policy |
| 10.1016/j.amepre.2023.09.004 | This knowledge can help strengthen intervention, prevention, and policy efforts aiming to mitigate the impacts of social adversities and trauma on persistent cardiometabolic health disparities over the lifecourse. | new evidence for policymaking |
| 10.2105/AJPH.2021.306599 | Our findings highlight the need to include LGBTQIA leaders and trusted individuals in the development of vaccination education and the delivery of vaccination services. | focus on specific subgroups |
| 10.1016/j.ypmed.2024.107970 | Conclusion: We discovered neighborhood social cohesion as an important obesogenic determinant that should be considered in policymaking to encourage higher levels of PA and higher diet quality. | new evidence for policymaking |
| 10.1016/j.amepre.2021.02.016 | Targeted efforts may be indicated to attenuate the risk and promote resilience among subgroups of young adults experiencing homelessness who are disproportionately affected by firearm violence. | alternative policy |
| 10.1016/j.ypmed.2023.107569 | We argue that these needs can be met with a person-centered, digital self-management core resource that supports people to better understand their needs and priorities and has links to find the resources they need to manage their health, alone or by judicious use of health services. | alternative policy |
| 10.2105/AJPH.2022.307160 | Conclusions. There is a paucity of PHSMS that measure individual-level racism, and few systems are linked to structural racism measures. Public Health Implications. Adopting a standard practice of racism surveillance can advance equity-centered public health praxis, inform policy, and foster greater accountability among public health practitioners, researchers, and decision-makers. Failure to explicitly address racism and the insufficient capacity to support a robust health equity data infrastructure severely impedes efforts to address and dismantle systemic racism. | alternative policy |
| 10.1093/eurpub/ckae103 | The regulations were in effect for different lengths of time and varied in some countries during the study period. The cumulative duration of MOC interruptions in all EU MS during the study period was 137 months (7.5% of the cumulative study period of 1836 months). Given the different approaches to the provision of MOCs in EU MS, it has proved appropriate to develop an optimal unified framework plan for future similar situations. | new evidence for policymaking |
| 10.1093/ije/dyae026 | It focuses then on three specific implications for the field: 1) promoting decoloniality in psychiatric epidemiology; 2) ensuring methodological rigor and feasibility; and 3) informing the development of mental health policy and services. | new evidence for policymaking |
| 10.1093/eurpub/ckaa139 | Therefore, policymakers should strengthen the quality of healthcare at primary care institutions and educate patients that these institutions are appropriate for managing chronic disease. | alternative policy |
| 10.1136/jech-2022-218794 | The use of multiple control groups strengthens the credibility of the results, making them useful for policy makers seeking solutions for universal health coverage. | new evidence for policymaking |
| 10.2105/AJPH.2022.307195 | Public health efforts to improve maternal health must address both access to care and universal screening for IPV. | alternative policy |
| 10.1016/j.ypmed.2021.106594 | National and regional disparities in environmental obesity determinants were identified that can inform targeted public health interventions. | new evidence for policymaking |
| 10.1136/jech-2022-219654 | Programme providers should target under-represented groups to ensure equitable access and narrow inequalities in T2DM. | focus on specific subgroups |
| 10.1016/j.ypmed.2022.107037 | Pandemic policies and recommendations should include and facilitate PA, specifically among vulnerable populations. | alternative policy |
| 10.1016/j.ypmed.2022.107186 | These results offer an important first systematic analysis of the trauma and mental health risks associated with community violence intervention practice and suggest that policymakers and practitioners should monitor and address worker risk of traumatic stress within this important public health profession. | focus on specific subgroups |
| 10.1093/eurpub/ckad199 | Discussion: Short-term exposure to outdoor air pollution may induce the occurrence or exacerbation of COPD patients; therefore, government departments should strengthen the management of air pollution, improve supervision and control mechanisms, pay attention to the quality of medical services, and reduce the adverse effects of air pollution on patients’ health. | alternative policy |
| 10.1016/j.amepre.2019.11.021 | Educational components should be added to interventions aimed to reduce food insecurity. Trial registration: This study is registered at www.clinicaltrials.gov NCT03492619. | alternative policy |
| 10.1016/j.amepre.2019.12.021 | Among male students, every category of violence victimization was more prevalent among gender-nonconforming than among gender-conforming students and most substance use categories demonstrated significant gender nonconformity disparities. After controlling for violence victimization, these disparities decreased but remained statistically significant for the use of cocaine (APR1=2.84 vs APR2=1.99), methamphetamine (APR1=4.47 vs APR2=2.86), heroin (APR1=4.55 vs APR2=2.96), and injection drug use (APR1=7.90 vs APR2=4.72). Conclusions: School-based substance use prevention programs may benefit from strategies that support gender diversity and reduce violence victimizations experienced by gender-nonconforming students, by providing a safe and supportive school environment. | focus on specific subgroups |
| 10.1093/aje/kwae084 | These results indicate that temperature vulnerability, particularly heat vulnerability, requires stronger public health and policy responses. This article is part of a Special Collection on Environmental Epidemiology. | new evidence for policymaking |
| 10.1016/j.ypmed.2022.107113 | This suggests that employer investments in diversity training and ally networks are effective interventions to enhance workplace culture, employee productivity and intergroup relations. | alternative policy |
| 10.1093/eurpub/ckac027 | Wherever not available, new national policies should enable coverage of travel and medical fees for living-donor surgery and follow-up for non-resident donors to improve uptake of LDKT in immigrant patients, and provide KT education that is culturally competent, individually tailored and easily understandable for patients and their potential living donors. | alternative policy |
| 10.1093/aje/kwab173 | For example, a 10-fold increase in gestational DAP concentration was associated with poorer longitudinally assessed Behavior Rating Inventory of Executive Function scores, as reported by mothers (β = 4.0 (95% confidence interval: 2.1, 5.8); a higher score indicates more problems), and Weschler Intelligence Scale for Children-Fourth Edition Working Memory scores (a 3.8-point reduction; â=.3.8 (95% confidence interval:.6.2, .1.3)).Reducing gestational exposure to OP pesticides through public health policy is an important goal. | new evidence for policymaking |
| 10.1136/jech-2020-215678 | This study identified independent groups with increased odds for seropositivity that may require specific surveillance measures to guide future protective interventions internationally, including vaccination once available. | focus on specific subgroups |
| 10.2105/AJPH.2024.307692 | Public health policy efforts need to be geographically tailored to address these disparities. | alternative policy |
| 10.1016/j.ypmed.2022.107240 | Transformative care models focusing on provider continuity, relationship building, and patient activation may offer more promise for improving birth outcomes than supplementing medical models with care management and other resources. | alternative policy |
| 10.1093/IJE/DYZ248 | Conclusions: These observational findings support policies to reduce mortality both through improving socio-economic circumstances and increasing education, and by altering intermediaries, such as lifestyle behaviours and morbidities. | new evidence for policymaking |
| 10.1016/S2468-2667(24)00179-8 | The data presented can assist policy makers and health authorities in mitigating increasing health inequalities by prioritising the protection of more susceptible areas and older population groups. We identify the projected areas of heightened risk (southern Europe), where policy intervention aimed at building adaptation and enhancing resilience should be prioritised. | new evidence for policymaking |
| 10.2105/AJPH.2020.306018 | Drawing these distinctions can help municipalities determine which immigrant-supportive measures are still permitted, and how best to mitigate the adverse public health effects of these preemption laws. (Am J Public Health. 2021;111:259-264. https://doi.org/10.2105/AJPH.2020.306018). | new evidence for policymaking |
| 10.1136/jech-2021-218074 | Conclusions Depression liability appears to cause increased non-employment, particularly by increasing disability. There was little evidence of depression affecting early retirement, hours worked or household income, but power was low. Effective treatment of depression might have important economic benefits to individuals and society. | new evidence for policymaking |
| 10.1136/jech-2020-214016 | There was no strong evidence for an association between SLI in childhood and other risk factors of cardiovascular disease. Conclusions Poor socio-economic conditions in childhood may contribute to the increased risk of premature cardiovascular disease among South Asians by raising their blood pressure. Elucidating the mechanisms and improving socio-economic conditions for children in South Asia could provide major reductions in the burden of cardiovascular disease. | alternative policy |
| 10.1093/ije/dyab101 | Conclusions: The wider impacts of pandemic mitigation strategies on non-COVID-19 infection-related hospitalizations are poorly understood. We observed marked and rapid decreases in hospitalized childhood infection. In tandem with broader consequences, sustainable measures, such as improved hand hygiene, could reduce the burden of severe childhood infection post-pandemic and the social and economic costs of hospitalization. | alternative policy |
| 10.2105/AJPH.2020.306096 | Wage-setting policies may be an important intervention for addressing risks of food insecurity among low-income workers. | alternative policy |
| 10.1093/eurpub/ckad076 | The factors identified in this systematic review should be considered as part of migrant-inclusive emergency preparedness strategies to address the disproportionate impact of health crises on migrant communities. | focus on specific subgroups |
| 10.1016/j.amepre.2021.04.008 | Enhanced screening and counseling using evidence-based practices during routine care for women with disabilities may be necessary to mitigate marijuana use. | alternative policy |
| 10.2105/AJPH.2023.307461 | Data from this analysis can inform occupational and public health research, policy, and interventions aimed at reducing the burden of disease and health inequities in the United States. (Am J Public Health. 2024;114(1):57–67. https://doi.org/10.2105/AJPH.2023.307461) | new evidence for policymaking |
| 10.1093/eurpub/ckae092 | Continued surveillance of the impact of service disruption on cancer services allows policy makers and strategic leaders in cancer control programmes to respond rapidly to mitigate the impact on cancer outcomes. | new evidence for policymaking |
| 10.1093/eurpub/ckae004 | Class 3 respondents (32%) were strongly reluctant to changes. Conclusions: Our study goes beyond average preferences and identifies three distinct population profiles, a majority open to reforms on specific aspects of care delivery, a smallest group in favour radical changes, and a third strongly against changes. Therefore, tailored approaches around healthcare reforms are needed, e.g. explaining the role of interprofessional teams in coordinating care, electronic health records and insurance premium variation. | alternative policy |
| 10.1136/jech-2020-213837 | Prescribed medications reinforced these findings; worsening crime rates were linked with antidepressant prescriptions among young stayers (OR=1.09; 95% CI 1.04 to 1.14) and with antipsychotic prescriptions among younger middle-aged movers (OR=1.11; 95% CI 1.01 to 1.23). Conclusion Changing neighbourhood crime exposure is related to individual mental health, but associations differ by psychiatric conditions, age and moving status. Crime reduction and prevention, especially in communities with rising crime rates, may benefit public mental health. | new evidence for policymaking |
| 10.1016/S2468-2667(24)00049-5 | To reduce the socioeconomic gap in life expectancy, effective efforts are needed to prevent early deaths from cardiovascular disease and cancer in socioeconomically deprived populations, with cancer prevention and control becoming an increasingly important field of action in this respect. Funding: German Cancer Aid and European Research Council. | focus on specific subgroups |
| 10.1016/j.ypmed.2021.106933 | Primary prevention strategies targeting these risk factors have the potential to drastically reduce stroke related morbidity and mortality. | alternative policy |
| 10.1016/j.amepre.2021.11.018 | The federal government should enact a national paid sick-leave law. | alternative policy |
| 10.1016/j.ypmed.2023.107769 | Policymakers must address all three inequalities and their fundamental causes. | alternative policy |
| 10.1016/S2468-2667(20)30146-8 | To reduce the socioeconomic gap in life expectancy, effective efforts are needed to prevent early deaths from cardiovascular disease and cancer in socioeconomically deprived populations, with cancer prevention and control becoming an increasingly important field of action in this respect. Funding: German Cancer Aid and European Research Council. | focus on specific subgroups |
| 10.1093/eurpub/ckad078 | These findings underscore the importance of targeted efforts to bring women with language barrier to prenatal care. | focus on specific subgroups |
| 10.1016/j.ypmed.2021.106906 | Conclusion: For reducing cervical cancer incidence and mortality, the readiness of health systems, the reach and effectiveness of new technologies and algorithms for increasing screening and treatment coverage, and the factors that support sustainability of these programmes need to be better understood. Answering these key IR questions could provide actionable guidance for countries seeking to implement the WHO Global Strategy towards cervical cancer elimination. | not a policy claim |
| 10.1016/j.amepre.2022.01.028 | Conclusions: Interventions enhancing consistent and nurturing parenting may help to reduce the long-term associations of neighborhood disadvantage with poor health. | alternative policy |
| 10.1093/eurpub/ckaa127 | Two major policy recommendations emerged: an urgent need for better health workforce data in Romania and development of more effective workforce management. | alternative policy |
| 10.1093/eurpub/ckz194 | The syndromic nature of long-term ill-health and functioning in ageing populations has implications for healthcare planning and public health policy in older populations. | new evidence for policymaking |
| 10.1016/j.ypmed.2020.106121 | Significant differences in the median rate of acute hepatitis B pre and post intervention in counties receiving vaccine were evaluated using Wilcoxon signed-rank test and bootstrapping. A Bland-Altman graph visualized significant differences in county-level rates of acute hepatitis B before and after the WV Pilot Project compared to the statewide estimate. Analyses identified significant geographic clustering of acute hepatitis B in southern WV across all four time-periods. Findings suggest that increased dissemination of hepatitis B vaccine through local health departments and existing harm reduction services can reduce the incidence of acute hepatitis B in states such as WV, which have been disproportionately affected by substance misuse. | alternative policy |
| 10.1016/j.amepre.2021.05.030 | Few studies of academic detailing for pre-exposure prophylaxis have been published to date; rigorous evaluation of HIV-specific adaptations and innovations of the approach would represent an important contribution. In the setting of the COVID-19 pandemic, interest in virtual delivery of academic detailing has grown, which could inform efforts to implement academic detailing in rural communities and other underserved areas. Increasing this capacity could make an important contribution to Ending the HIV Epidemic in the U.S. and other HIV prevention efforts. | alternative policy |
| 10.1136/jech-2022-219095 | Conclusions Both the heterogeneity among caregivers and the related contextual factors should be accounted for by policymakers as well as in future research investigating the health impact of informal caregiving. | focus on specific subgroups |
| 10.1136/jech-2023-221606 | Conclusions Our findings imply that the effect of retirement should be considered within a cultural context to inform suitable and effective strategies to alleviate loneliness. | new evidence for policymaking |

**Supplementary Table 4. Policy claims (after removal of 1059 potential methodological papers (2.3%))**

|  | **1990-1999** | **2000-2009** | **2010-2019** | **2020-2024** | **All years** |
| --- | --- | --- | --- | --- | --- |
| Number of articles, N | 10014 | 12266 | 15241 | 7227 | 44748 |
| Number of unique journals, N | 9 | 9 | 10 | 10 | 10 |
| Words in abstracts, mean (SD) | 206.0 (67.9) | 216.2 (55.4) | 221.2 (52.1) | 238.9 (57.4) | 219.3 (58.6) |
| Papers with keywords included, % | 47.4 | 59.8 | 44.4 | 57.0 | 51.3 |
| Countries of corresponding author, N | 112 | 118 | 121 | 104 | 153 |
| Policy claims, N (%) | 1825 (18.2) | 2844 (23.2) | 4362 (28.6) | 2628 (36.4) | 11659 (26.1) |
| All journals/countries | 18.2 | 23.2 | 28.6 | 36.4 | 26.1 |
| Journals |  |  |  |  |  |
| Epidemiology | 1.2 | 4.3 | 4.6 | 5.7 | 3.9 |
| European Journal Of Epidemiology | 19.3 | 15.4 | 10.0 | 14.3 | 14.5 |
| American Journal Of Epidemiology | 8.7 | 9.0 | 11.5 | 16.4 | 10.4 |
| International Journal Of Epidemiology | 15.3 | 20.1 | 16.4 | 20.5 | 17.5 |
| Journal Of Epidemiology And Community Health | 20.8 | 25.9 | 30.4 | 41.3 | 28.8 |
| European Journal Of Public Health | 28.2 | 33.6 | 40.4 | 45.2 | 38.7 |
| Preventive Medicine | 25.1 | 30.3 | 38.0 | 45.3 | 35.7 |
| American Journal Of Public Health | 24.6 | 33.7 | 38.5 | 46.2 | 33.6 |
| American Journal Of Preventive Medicine | 31.4 | 37.2 | 35.8 | 47.2 | 38.1 |
| The Lancet Public Health | N/A | N/A | 58.7 | 64.1 | 62.4 |
| Countries |  |  |  |  |  |
| Norway | 8.8 | 6.4 | 14.5 | 16.9 | 11.0 |
| Japan | 9.2 | 17.4 | 17.7 | 17.8 | 16.0 |
| Denmark | 12.9 | 9.2 | 11.5 | 25.4 | 13.6 |
| Sweden | 14.3 | 15.3 | 22.2 | 27.4 | 18.9 |
| Germany | 22.5 | 18.6 | 16.7 | 29.8 | 20.5 |
| Finland | 10.7 | 18.7 | 25.8 | 30.5 | 20.3 |
| China | 11.4 | 17.2 | 23.0 | 33.3 | 25.8 |
| United kingdom | 16.8 | 23.4 | 25.6 | 34.5 | 24.9 |
| Netherlands | 15.0 | 22.3 | 25.7 | 34.5 | 23.2 |
| Spain | 15.8 | 25.1 | 26.8 | 35.1 | 25.5 |
| Canada | 17.8 | 23.7 | 29.9 | 35.3 | 27.5 |
| France | 17.5 | 20.4 | 25.7 | 35.9 | 24.0 |
| United states | 18.9 | 24.6 | 30.8 | 38.3 | 27.5 |
| Australia | 24.3 | 26.9 | 30.0 | 41.1 | 30.3 |
| Italy | 15.2 | 19.8 | 36.2 | 43.5 | 23.9 |
